# Genomic privacy risks in GWAS summary statistics

**DOI:** 10.1101/2025.09.09.25335252

**Authors:** Ao Lan, Yudi Pawitan, Xia Shen

**Affiliations:** Biostatistics Group, School of Life Sciences, Sun Yat-sen University, Guangzhou, China; Department of Medical Epidemiology and Biostatistics, Karolinska Institutet, Stockholm, Sweden; Center for Intelligent Medicine Research, Greater Bay Area Institute of Precision Medicine (Guangzhou), State Key Laboratory of Genetic Engineering, Center for Evolutionary Biology, School of Life Sciences, Fudan University, Shanghai, China; Centre for Global Health Research, Usher Institute, University of Edinburgh, Edinburgh, UK

## Abstract

The rapid advancement in sequencing technologies has exponentially increased the availability of genomic data, heightening concerns about data privacy. Despite the perceived safety of publicly accessible genome-wide association study (GWAS) summary statistics, we demonstrate that their combination with less sensitive high-dimensional phenotype data can lead to significant leakage of confidential genomic information. By transforming a linear regression model into linear programming constraints, we scrutinize the potential for genomic data recovery using GWAS summary statistics. We found that an effective phenotype-to-sample size ratio above 0.85 could enable full genotype recovery, and that above 0.16 was sufficient to enable individual identification. Certain non-European populations are especially vulnerable. The results stress the urgent need for stronger privacy protections in genomic research while maintaining data utility.

## Introduction

The unprecedented growth in genomic data, driven by advances in sequencing technology, has brought data privacy issues to the forefront of genomic research. Large-scale initiatives such as the GTEx project, the 1000 Genomes Project, the UK Biobank, and the All of Us Research Program exemplify this surge in genomic data generation (***GTEx Consortium, 2020; 1000 Genomes Project Consortium, 2015; Bycroft et al., 2018; All of Us Research Program Investigators, 2019***). With this influx, addressing privacy concerns becomes critical (***Bonomi et al., 2020; Erlich and Narayanan, 2014; Lunshof et al., 2008***).

Genomic data’s sensitivity is highlighted by the restricted access to whole-genome sequencing (WGS) data in projects like GTEx, contrasting with the open accessibility of phenotype data, including RNA-seq read counts. Similarly, human genetics cohorts, such as the INTERVAL study on proteogenomics, share their high-throughput proteomics data, while access to their genomic data is restricted (***Sun et al., 2018***). UK Biobank projects typically require specific applications to access genomic data. Conventionally in human genetics, genome-wide association studies (GWAS) summary statistics are often publicly shared, presumed safe under the assumption that individual genotypes cannot be easily inferred (***Pasaniuc and Price, 2017a***). However, emerging evidence suggests that this assumption is flawed, with even non-sensitive data posing significant privacy risks.

Recent work by Walker et al. highlights how individuals in single-cell gene expression datasets are vulnerable to linking attacks, despite the inherent noise in single-cell measurements (***Walker et al., 2024***). This study demonstrated that publicly available cell-type-specific eQTL information could be exploited to infer sensitive phenotypic or genotypic information. Furthermore, the authors developed methods for genotype prediction and genotype-phenotype linking that remain effective even without relying on eQTL data. These findings underscore the growing privacy concerns surrounding publicly available data, revealing that even datasets traditionally considered non-sensitive, such as gene expression or cell-type-specific profiles, can lead to private information leakage when combined with publicly available resources.

In GWAS, linear regression models are commonly used to estimate the marginal effects of genetic variants on traits. Here, we show that as the number of phenotypes grows, the summary association statistics provide sufficient linear programming (LP) constraints, enabling the recovery of the genotype data of each genetic variant. This study extends prior work by focusing on the risks posed by GWAS summary statistics, particularly when combined with high-dimensional phenotype data such as omics traits. Our findings reveal that the ratio of the effective number of phenotypes to the sample size (*R/N*) plays a pivotal role in recovery accuracy. Additionally, we demonstrate that genetic variants with lower minor allele frequencies (MAF) are more susceptible to recovery, an observation consistent across both simulations and real genomic datasets.

By incorporating theoretical insights, simulation-based validations, and applications to real-world datasets, this study provides a comprehensive framework for understanding and mitigating the risks of private information leakage in genomic research. The results underscore the urgent need for enhanced privacy safeguards to ensure a balance between data utility and privacy protection.

## Results

### Summary association statistics of high-dimensional phenotypes enables genotypic data recovery

The theoretical basis for recovering genotype data for a genetic variant when the number of phenotypes with available summary association statistics is sufficiently large revolves around the use of an LP approach. Starting from the linear model in GWAS, where the phenotype matrix **Y** for *N* samples and *T* traits is modeled based on the genotype **X**_*j*_ of variant *j* and an error term *ϵ*_*jt*_. Through least squares estimation, the marginal effect size 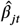 and its standard error 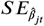 are expressed as functions of **X**_*j*_, the genotype, and **Y**_*t*_, the phenotypic data. The equation derived for 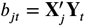 is used to establish constraints for an LP model.

The key idea is that with a large number of traits *T*, the association statistics *b*_*jt*_ provide multiple independent constraints on **X**_*j*_, allowing accurate inference of the genotype. Due to numerical precision issues, the exact constraint cannot always yield feasible solutions. Nevertheless, the constraint can be relaxed to an interval approximation, and an LP solver can be used to estimate 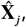 rounded to integer values that correspond to allele counts.

Genomic data recovery hinges on the information captured by the *N* × *M* phenotype matrix, where additional dependent traits do not contribute new information. The rank of the phenotype matrix (*R*) serves as a metric for the effective number of independent traits. When *R* is sufficiently large relative to *N*, the information becomes saturated to recover the genotype data (see **Methods and Materials**). We investigated the relationship between the *R/N* ratio and the accuracy of genotype data recovery.

Simulations revealed a strong correlation between recovery accuracy and the *R/N* ratio (**Figure 1**). Notably, sample size *N* appeared independent of recovery accuracy as long as the *R/N* ratio remained constant. Our simulations indicate that an *R/N* ratio of 0.85 or higher ensures complete genotype recovery (**Figure 1**).

**Figure 1.**
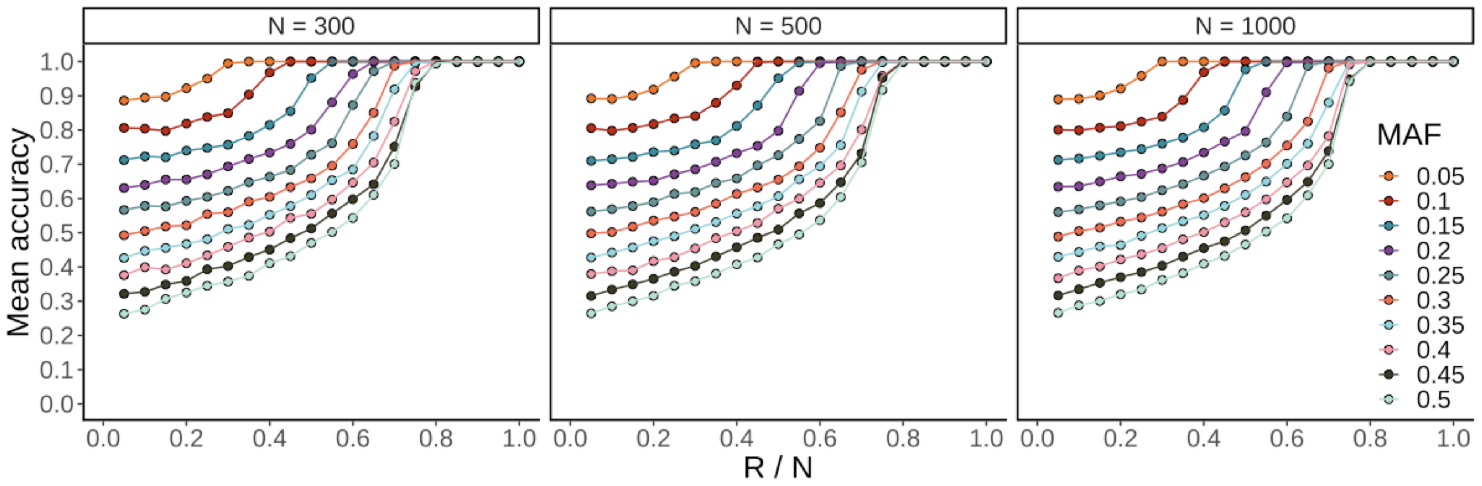
Theoretical genotype recovery accuracy in relation to sample size, *R/N* ratio, and MAF. Simulations were performed for sample sizes of 300, 500, and 1000, with *R/N* ratios ranging from 0.05 to 1 in increments of 0.05 and MAFs ranging from 0.05 to 0.5. Each point represents the mean accuracy of 30 replications.

### Simulation based on 1000 Genomes Project data

Using the genotype data of 1 million single nucleotide polymorphisms (SNPs) for 489 European ancestry samples in the 1000 Genomes Project (https://www.internationalgenome.org), we simulated the phenotypic data for 4,890 (2 × 489 × 5) independent phenotypes based on a linear model, with five different heritability settings (**Methods and Materials**). A standard single-phenotype GWAS analysis was conducted for each simulated phenotype to obtain the summary association statistics. Subsequently, 10,000 SNPs were randomly selected for each MAF bin to assess the genotype recovery performance under different *R/N* ratios and heritability scenarios (**Figure 2**).

**Figure 2.**
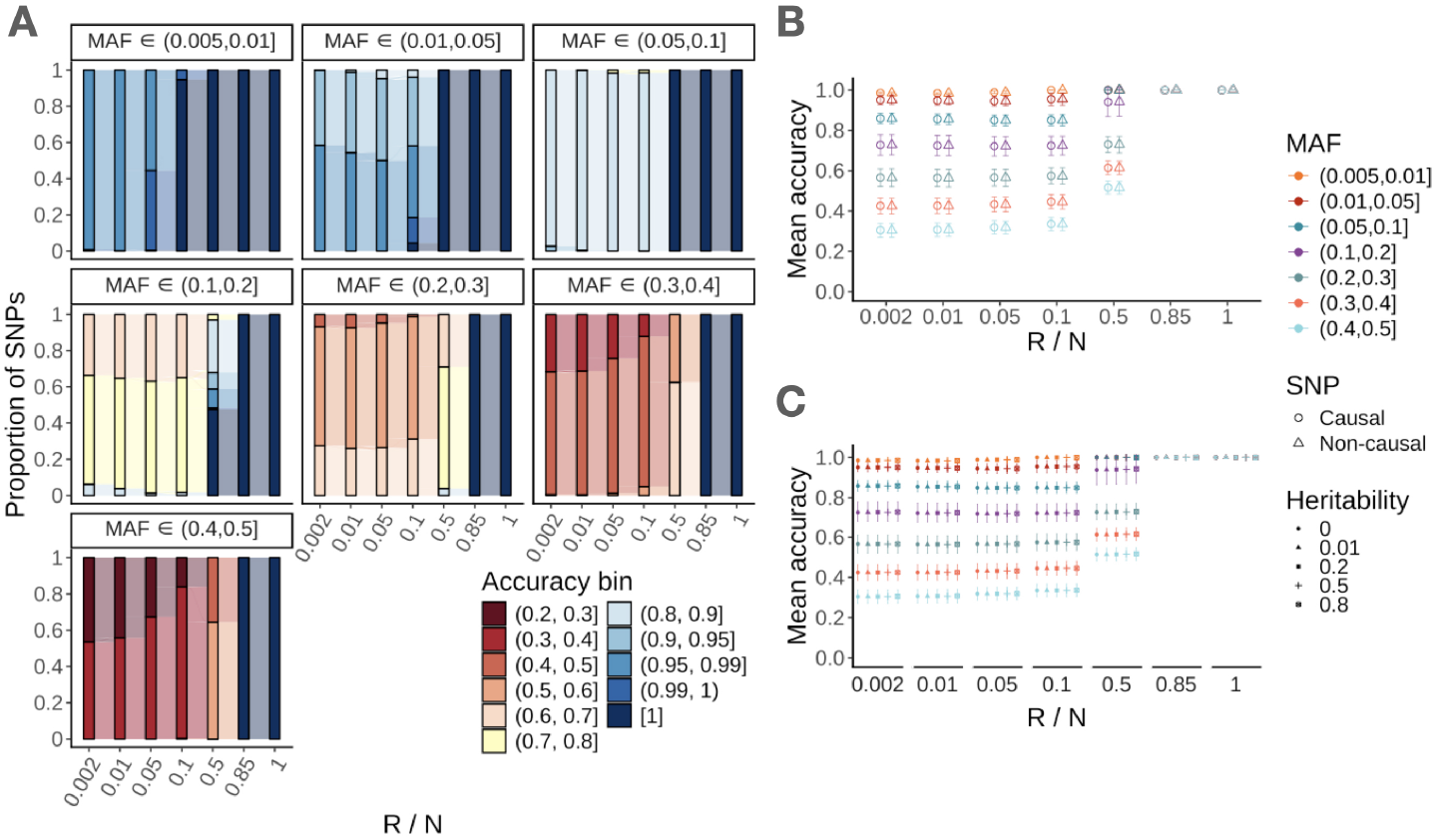
Recovery performance in simulations based on 489 European samples from 1000 Genomes Project. Genotypic data of 1 million SNPs were used in the simulation. Five replications were conducted for each *R/N* ratio to obtain the average recovery accuracy of each SNP. **(A)** depicts simulations with no causal SNPs (heritability = 0). **(B)** shows the mean accuracy of each MAF bin, contrasting causal and non-causal SNPs at heritability = 0.5. **(C)** presents the mean accuracy across various MAF bins, *R/N* ratios, and heritability levels. In simulations with heritability > 0, 10% of randomly selected SNPs were designated as causal. Error bars represent standard deviations.

As expected from the theory, the simulation confirmed that the genotypes for SNPs with lower MAFs could be recovered with smaller *R/N* ratios. Real genomic data simulations, dividing SNPs into MAF bins, support this finding. Genotypes for SNPs with MAF *<* 0.1 could be completely recovered if *R/N >* 0.5. More common SNPs (MAF *>* 0.1) require an *R/N* ratio of 0.85 or above to fully recover their genotypic data.

As the LP solution for the genotypic data is mathematical, it is independent of the genotype-phenotype map. The simulation also showed that the recovery accuracy of the genotypic data was independent of the SNP’s causality nor the heritability of the phenotype (**Figure 2B, C**).

### The risk of personal genomic data leakage in the GTEx project

As an example, the open-access data of the whole blood tissue from the GTEx project include cis-region GWAS summary statistics of the expression quantitative trait loci (eQTL) analysis, the original gene expression phenotypes, and the corresponding covariate data in the GWAS analysis (***GTEx Consortium, 2020***). The overlapping cis-regions led to multiple traits (expression levels of different genes) having summary association statistics at the same variants. The distribution of the number of variants across MAFs and *P /N* ratios (where *P* is the number of traits, *N* = 369) is shown in **Figure 3A**.

**Figure 3.**
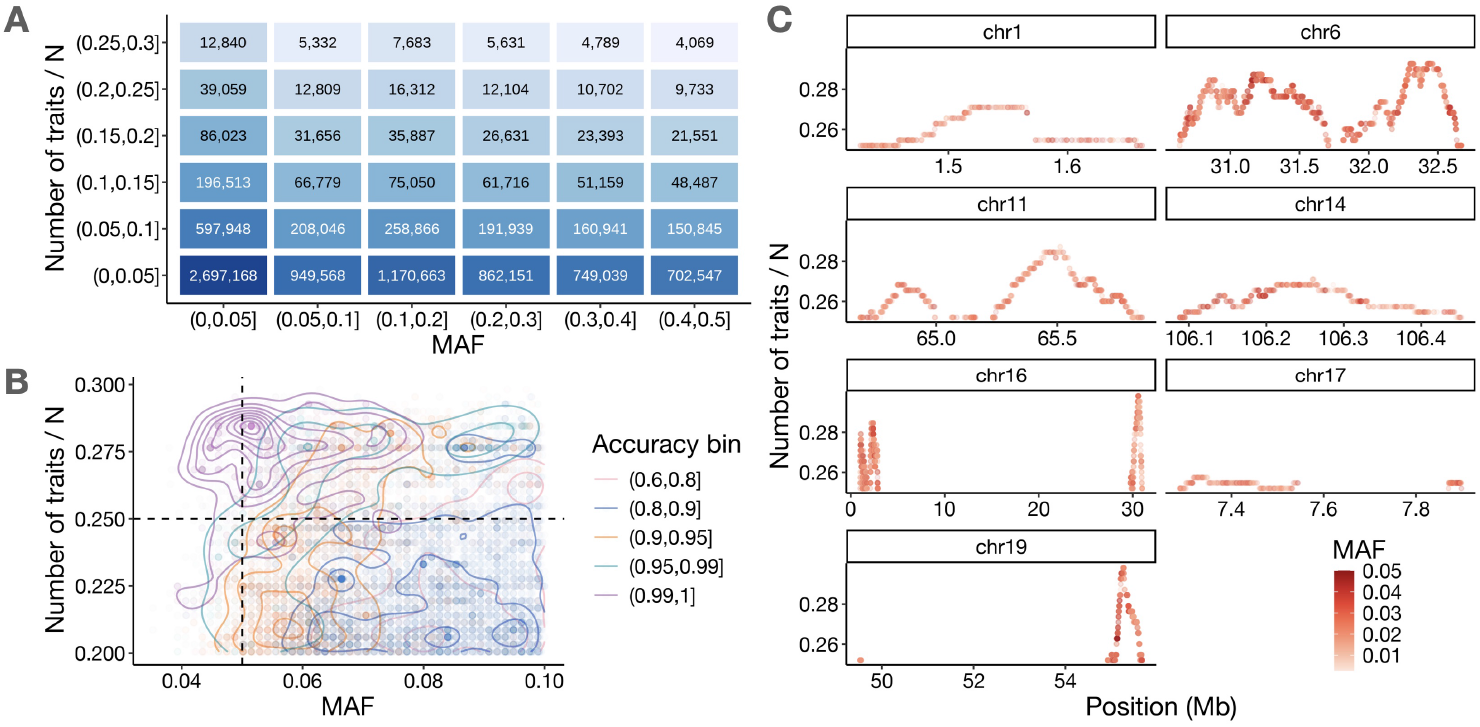
Recovery application in GTEx. The algorithm was applied to cis-region GWAS summary statistics for 20,059 genes from GTEx v7 whole blood tissue, with open-access phenotype and covariate data. Due to overlapping cis-regions, some variants had GWAS data for multiple traits (genes). **(A)** shows the distribution of variants across MAF and the number of traits (P) divided by the sample size (N = 369). **(B)** illustrates the recovery accuracy for 17,568 variants (MAF ≤ 0.1, *P /N* > 0.2) with true genotype. The vertical and horizontal lines represent x = 0.05 and y = 0.25, respectively. **(C)** shows the genomic positions (GRCh37) of 12,840 variants with MAF ≤ 0.05 and *P /N* > 0.25.

Recovery accuracy was assessed by comparing the recovered genotype data to the true genotype data of 17,568 overlapped variants (MAF ≤ 0.1 and *P /N* > 0.2) from ***Pan et al. (2023***). 93.28% variants had recovery accuracy greater than 80%, where 44.36% variants had an accuracy greater than 90%. 36.08% individuals had perfectly recovered genotypic information for at least 90% genetic variants.

Specifically, for the 346 variants with MAF ≤ 0.05 and *P /N* > 0.25, the genotype data recovery accuracy was greater than 99% (**Figure 3B**). By assuming MAF ≤ 0.05 and *P /N >* 0.25 as criteria for “easily recoverable” variants, we examined the genomic positions (GRCh37) of all the 12,840 “easily recoverable” variants. The genomic regions with a high risk of genotypic data leakage are identified in **Figure 3C**. These results demonstrate the potential for genomic data leakage for the GTEx samples.

### Risk assessment for accurate individual identification in different ancestries

In practice, the key risk of leaking genomic information is that it may allow the identification of a specific individual in a population. Here, we sought to assess the *R/N* ratio threshold required to identify specific samples based on the GWAS summary information of a set of SNPs. According to our simulations (*N* = 1, 000), we obtained an empirical function that determines genotype recovery accuracy based on two variables, MAF and *R/N* ratio (**Figure 4A**). We fitted a single-parameter curve, *R/N* = 2*/*(1 +exp{*b* × MAF}) − 1 to best separate the scenarios of genotype recovery accuracy greater than 0.99 and less than or equal to 0.99. We obtained a least-square estimate of 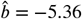 (standard error 0.39), giving a fitted lower bound for genotype recovery accuracy greater than 0.99.

**Figure 4.**
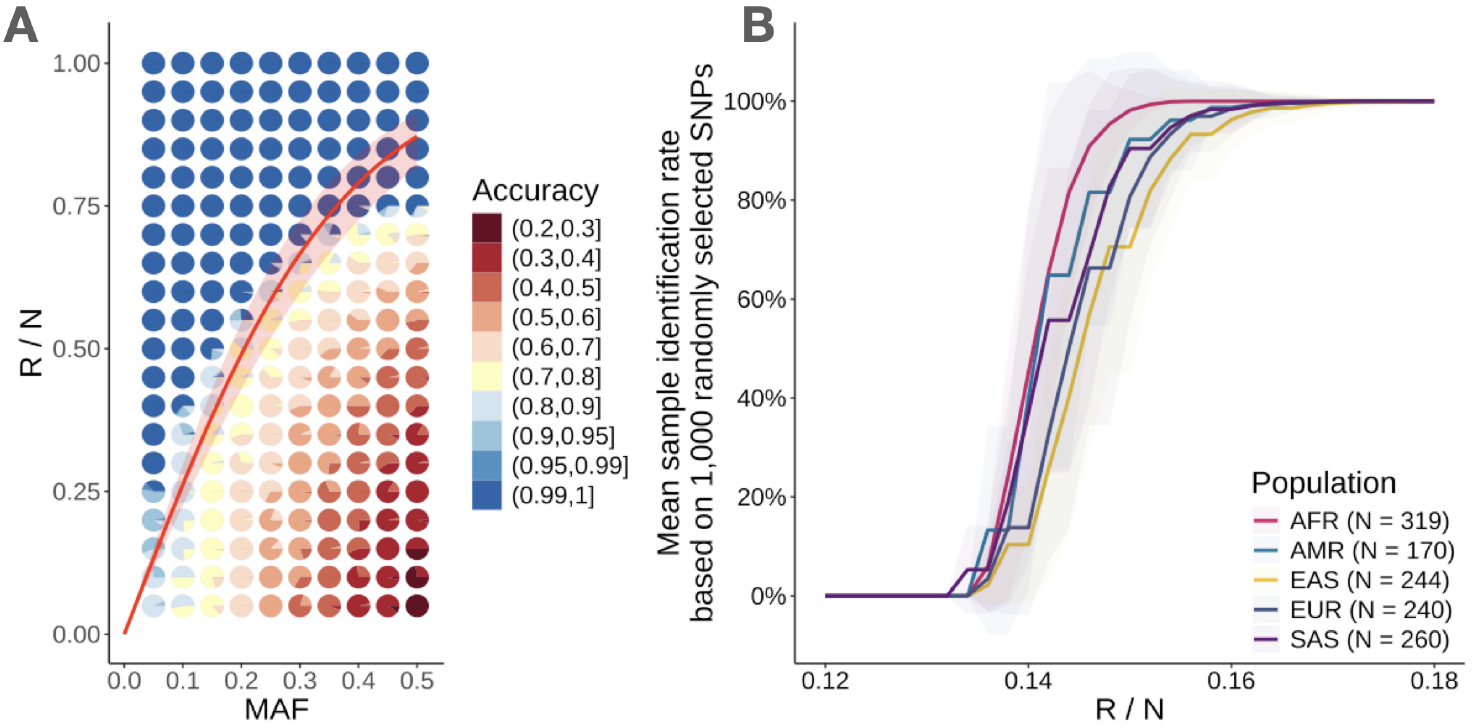
Minimal *R/N* ratio predicted by MAF for sample identification. The boundary curve separating recovery accuracy > 0.99 from accuracy ≤ 0.99 was fitted based on theoretical simulations of 1,000 samples, as shown in **Figure 1**. The model for this boundary is described by the equation *R/N* = 2 ÷ (1 + *e*^*b*×MAF^) − 1, where *b* is the parameter to be estimated. **(A)** The red line represents the fitted curve based on points at the lower boundary of accuracy > 0.99. **(B)** Displays the mean sample identification rate of 30 replications based on 1,000 randomly selected common SNPs (MAF > 5%) along with the corresponding *R/N* cutoff. For a given *R/N* cutoff, only SNPs with predicted accuracy > 0.99 are retained for sample identification in populations from the 1000 Genomes Project (EUR: European, AFR: African, AMR: Admixed American, EAS: East Asian, SAS: South Asian). Shadings are 95% confidence intervals.

Subsequently, for individual-level data from different ancestries of the 1000 Genomes Project, we randomly selected 1,000 common SNPs (MAF > 5%) and assessed the sample identification rate, i.e., the proportion of individuals that can be identified using the genotype of “leaked” SNPs whose MAF-predicted accuracy was greater than 0.99 (**Figure 4B**). We found that, based on 1,000 common varints, an *R/N* ratio greater than 0.16 was sufficient to identify nearly all the individuals. African population appeared to have higher risks, as common SNPs mostly identified in other ethnic groups tend to have lower MAF among the Africans. Note that an *R/N* ratio of 0.16 is much lower than the value 0.85 we demonstrated above for complete genotype recovery of a SNP, as it is not required to perfectly recovery all the genotype data for 1,000 SNPs in order to identify a specific individual from the rest of the population.

## Discussion

The perceived safety of publicly available GWAS summary statistics is critically challenged when combined with additional phenotype data, potentially exposing sensitive genomic information. Our study demonstrates that by using linear programming constraints derived from GWAS summary statistics, genotype data for individual genetic variants can be recovered with high accuracy when the ratio of effective traits to sample size (*R/N*) exceeds a critical threshold. This highlights a major vulnerability in current data-sharing practices, emphasizing the need for more robust privacy safeguards in genomic research.

With the increasing prevalence of high-dimensional omics datasets, such as proteomics, transcriptomics, and metabolomics, the number of traits available for GWAS analyses has grown exponentially (***Sun et al., 2018; GTEx Consortium, 2020***). This abundance of phenotypic data exacerbates the risk of genomic data leakage, as more traits effectively increase the constraints available for recovering individual-level genotype data. Our findings underscore the importance of the *R/N* ratio, with simulations demonstrating that a ratio exceeding 0.85 is sufficient for complete genotype recovery of SNPs, particularly for those with low minor allele frequencies (MAF).

This privacy concern aligns with earlier work highlighting vulnerabilities in genomic data-sharing practices. Erlich and Narayanan (2014) showed that even anonymized genomic data could be reidentified by combining public records and genetic genealogy (***Erlich and Narayanan, 2014***). Similar risks arise from phenotypic data sharing, as seen in the GTEx project, where transcriptomic data and GWAS summary statistics could potentially be exploited for genomic recovery. Our results extend this line of research by demonstrating that recovery risks are not limited to genotypic reidentification but can also enable the full recovery of genotype matrices for entire cohorts.

Our analysis highlights the critical role of MAF in determining recovery accuracy. Genetic variants with low MAF are more susceptible to recovery, requiring smaller *R/N* ratios for complete recovery. This aligns with previous findings on the vulnerability of rare variants to imputation and recovery methods (***Pasaniuc and Price, 2017b***). Additionally, we observed that recovery accuracy is largely independent of SNP heritability or causality, suggesting that privacy risks are not mitigated by excluding specific classes of SNPs. Instead, the intrinsic characteristics of the genotypic data, particularly sparsity under Hardy-Weinberg equilibrium, drive recovery feasibility.

Our study assumes the use of conventional computational techniques for genotype recovery. However, emerging technologies such as quantum computing could significantly amplify these risks. Quantum algorithms are inherently well-suited for solving optimization problems, including those involving linear programming constraints. Appendix 2 illustrates that with future computational advancements, such as quantum annealing, genotype recovery could become feasible even with fewer traits and lower *R/N* ratios. This raises the alarming possibility that a cohort’s genotypes could be fully reconstructed using only one or a few traits’ GWAS summary statistics, rendering even low-dimensional phenotypic data a potential threat to genomic privacy. Such scenarios necessitate a forward-looking approach to data protection, accounting for technological advancements that could render current safeguards obsolete.

While our findings emphasize the privacy risks of GWAS summary statistics, they also highlight the need for careful consideration of trade-offs between data utility and privacy. The sharing of GWAS summary statistics has been instrumental in advancing our understanding of complex traits and diseases, enabling meta-analyses and the development of polygenic risk scores (***Visscher et al., 2017; Torkamani et al., 2018***). However, the potential for genomic data leakage calls for stricter data governance policies, such as controlled access repositories, encryption techniques, and differential privacy methods (***Dwork, 2006***).

Several strategies have been proposed to mitigate privacy risks while maintaining data utility. For instance, differential privacy methods can introduce noise into summary statistics to prevent individual-level inferences while preserving aggregate statistical properties (***Yu et al., 2014***). Additionally, federated learning approaches, which enable collaborative analyses without sharing raw data, hold promise for reducing privacy risks in genomic research (***Rieke et al., 2020***).

Our findings have significant implications for genomic data-sharing policies. Large-scale initiatives such as the UK Biobank and the GTEx project should reassess their data-sharing practices, particularly for phenotypic data with high dimensionality. Implementing stricter access controls, auditing data-sharing agreements, and adopting advanced privacy-preserving techniques could help mitigate the risks identified in this study.

Future research should explore the impact of incorporating advanced computational methods, such as quantum algorithms and machine learning, into genotype recovery models. These methods could reduce computational barriers, enabling more efficient genotype reconstruction and necessitating even stricter safeguards. Additionally, further investigations into the interplay between phenotypic dimensionality, sample size, and recovery accuracy could inform the development of guidelines for safe data-sharing practices in genomic research.

Our study demonstrates that publicly available GWAS summary statistics, when combined with high-dimensional phenotypic data, pose a significant risk to genomic privacy. The *R/N* ratio and MAF are critical factors influencing recovery accuracy, with low MAF SNPs being particularly vulnerable. Emerging computational techniques, such as quantum computing, could further exacerbate these risks, underscoring the urgency of implementing robust privacy safeguards. By elucidating the factors influencing genotype recovery, this study contributes to the ongoing efforts to balance data utility and privacy in genomic research.

## Methods and Materials

### Summary association statistics as constraints for linear programming

The linear model used in GWAS assumes that the phenotype matrix **Y** for *N* samples and *T* traits has the form of **Y**_*t*_ = **X**_*j*_*β*_*jt*_+***ϵ***_*jt*_, where **X**_*j*_ is the genotype vector of SNP *j*, and ***ϵ***_*jt*_ is the corresponding error term. The least squares estimate of *β*_*jt*_ is 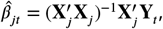, with standard error from:

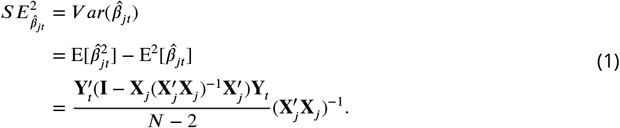

Denoting 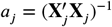 and 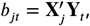, we have

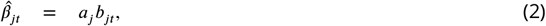

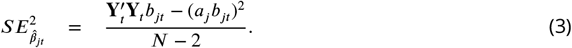

By solving the equations (2) and (3), we obtain the following equation:

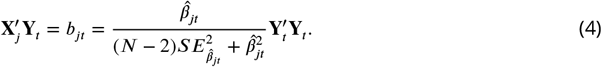

Equation (4) serves as a linear programming (LP) constraint to solve for the genotype data of SNP *j*. Numerical precision can lead to no feasible solution when the constraint is applied in LP. To solve this issue, we relax the constraint in eq. (4) as follows:

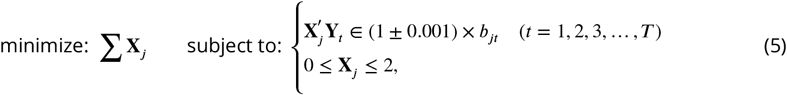

With sufficiently large *T*, we estimate 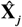 using the *linprog* package in R ***R Core Team (2021***). In theory, because the genotype data are a vector of allele counts as 0, 1, and 2, we should constrain **X**_*j*_ ∈ ℤ^*n*^ and apply integer programming, which is computationally ineffective. For fast computation, we neglect the integer constraint to solve for allelic dosages, and we round 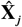 into 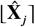 with elements replaced by their closest integers.

The validity of this linear programming formulation relies on the assumption that the GWAS marginal effect estimates 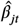 across phenotypes *t* are statistically independent when the phenotypes themselves are independent. In Appendix 1, we prove that if two phenotypes *Y*_1_ and *Y*_2_ are independent, then their corresponding genetic effect estimates 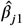 and 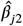 are also independent for any given SNP *j*. This ensures that each phenotype contributes an independent constraint to the linear programming system, and the number of independent traits (i.e., the rank *R* of the phenotype matrix) reflects the information available for genotype recovery.

### Simulations

For the theoretical simulations, we draw independent diploid genotypes of N = 300, 500, and 1,000 samples from a binomial distribution ***B***(2, MAF), with MAF ranging from 0.05 to 0.5 in increments of 0.05. For each specific *R/N* ratio, which varied from 0.05 to 1 in increments of 0.05, we simulated *R* independent traits from a standard normal distribution. We summarized the resulting recovery accuracies from 30 independent replicates of each parameter scenario. Recovery accuracy for each SNP was defined as the proportion of correctly recovered genotypes, i.e., the number of correctly recovered samples divided by the total number of samples.

To investigate various scenarios in real genomic data, we utilized the *N* = 489 European samples from the 1000 Genomes Project Phase 3 dataset (***1000 Genomes Project Consortium, 2015***), obtained from the LD Score Regression reference files website (https://alkesgroup.broadinstitute.org/LDSCORE/). To align with the quality control on samples of GWAS studies, we also checked the kinship using the KING (v2.2.1) (***Manichaikul et al., 2010***) to ensure that samples were unrelated. The dataset contained 9,997,231 SNPs with MAF greater than 0.5%. Without losing generality, we randomly selected 1,000,000 SNPs with PLINK (v1.90b6.5) (***Purcell et al., 2007***) to perform GWAS analysis. **1)** A total of 978 (2*N*) traits were simulated from a standard normal distribution for the 489 samples, i.e., no genetic effect. Following this, *R* traits were sampled from the 978 traits based on a predetermined *R/N* ratio, with values of 1/N, 0.01, 0.05, 0.1, 0.5, 0.85, or 1.0. Next, we randomly selected 10,000 SNPs for MAF bins of 0-0.005, 0.005-0.01, 0.01-0.05, 0.05-0.1, 0.1-0.2, 0.2-0.3, 0.3-0.4, and 0.4-0.5 to evaluate the recovery performance under different *R/N* ratios. For each *R/N* ratio, we replicated the analysis five times to obtain a mean recovery accuracy for each SNP. **2)** For GWAS simulations with heritabilities of 0.01, 0.2, 0.5, and 0.8, we randomly selected a set of 10% of the SNPs as causal with effect sizes drawn from a normal distribution and repeated the steps in **1)**.

### GTEx data analysis

Data for the whole blood tissue were obtained from the GTEx portal (https://gtexportal.org/, version phs000424.v7.p2.c1). This dataset includes open-access cis-region eQTL GWAS summary statistics (9,565,634 variants), transcriptome-wide expression data (369 samples), and the covariates used in the eQTL GWAS analysis (369 samples). The genotype data of 6,496,708 variants and 373 samples are restricted as sensitive information, and access can only be granted via approved projects from the dbGaP repository under accession phs000424.v7.p1.

The overlapping sample size between the expression phenotype data and genotype data was 352, while the number of overlapping variants between the GWAS and genotype data was 1,228,905. From this set of overlapping variants, we performed genotype recovery on a subset of 17,568 variants with MAF ≤ 0.1 and *P /N >* 0.2, where *P* represents the number of traits (in this case genes) and *N* = 369. The MAFs of the variants were derived from the real genotype data.

### Predicted minimal *R/N* for sample identification

The boundary curve separating recovery accuracy greater than 0.99 from accuracy less than or equal to 0.99 was estimated using theoretical simulations of 1,000 samples. To ensure that the curve intersects the origin (i.e., when MAF = 0 and *R/N* = 0), we modeled the boundary curve using the equation *R/N* = 2*/*(1 + exp{*b* × MAF}) − 1, where *b* is a single parameter to be estimated. Data points located at the lower boundary of accuracy greater than 0.99 were used to fit the curve. We then randomly selected 1,000 common SNPs (MAF *>* 5%) from the 1000 Genomes Project data and calculated the predicted recovery accuracy for each SNP based on the *R/N* ratio. We then identified SNPs with predicted accuracy greater than 0.99 and used them to estimate the sample identification rate across different populations, i.e., the proportion of samples in the population that can be distinguished from others, calculated based on the genotypes of the “leaked” SNPs. For each population ancestry, we conducted 30 replicates to obtain a mean sample identification rate and a 95% confidence interval.

## Data Availability

1000 Genomes Project Phase 3 data are available at: https://www.internationalgenome.org/, and the curated data were obtained from https://alkesgroup.broadinstitute.org/LDSCORE/. The GTEx Project data are available at: https://gtexportal.org/home/. The individual genotype data used for validation analysis were obtained from the dbGaP repository under accession phs000424.v7.p2.c1 and phs000424.v7.p1, respectively.

https://github.com/now2014/solveX

## Data and code availability

1000 Genomes Project Phase 3 data are available at: https://www.internationalgenome.org/, and the curated data were obtained from https://alkesgroup.broadinstitute.org/LDSCORE/. The GTEx Project data are available at: https://gtexportal.org/home/. The individual genotype data used for validation analysis were obtained from the dbGaP repository under accession phs000424.v7.p2.c1 and phs000424.v7.p1, respectively. All original code has been deposited on GitHub (https://github.com/now2014/solveX).

## Acknowledgments

X.S. was in receipt of a National Key Research and Development Program grant (No. 2022YFF1202105), a National Natural Science Foundation of China (NSFC) grant (No. 12171495), and a Swedish Research Council (Vetenskapsrådet) grant (No. 2022-01309). We thank the 1000 Genomes Project Consortium and the GTEx Consortium for providing the individual-level data.

## Additional information

### Author contributions

X.S. conceived the project and supervised the study. A.L. and X.S. developed the method. A.L. performed data analysis. Y.P. assisted with the validation analysis. All authors wrote and reviewed the manuscript.

## Appendix 1. Proof of the independence of genetic effects estimates for independent phenotypes

Let *Y*_1_ and *Y*_2_ be independent random variables, and let *X* be another random variable. Here we show that the regression slopes of *Y*_1_ on *X* and *Y*_2_ on *X* are also independent. The regression slope of *Y*_1_ on *X*, denoted by 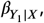 is given by 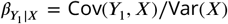. Similarly, the regression slope of *Y*_2_ on *X*, denoted by 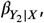 is given by 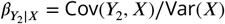. Since *Y*_1_ and *Y*_2_ are independent, we have Cov(*Y*_1_, *Y*_2_) = 0. Consider the covariance between the slopes 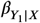 and 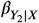:

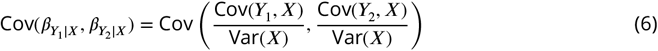

Since Var(*X*) is a constant, it can be factored out of the covariance:

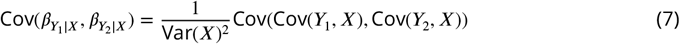

To proceed, we use the bi-linearity of covariance:

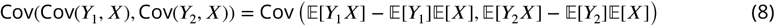

Given the independence of *Y*_1_ and *Y*_2_, and noting that the covariance between expectations of independent variables is zero, we simplify:

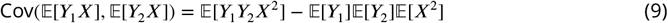

Since *Y*_1_ and *Y*_2_ are independent, 𝔼 [*Y*_1_*Y*_2_] = 𝔼 [*Y*_1_] 𝔼 [*Y*_2_], thus:

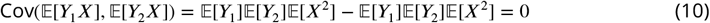

Therefore, the covariance between the slopes is:

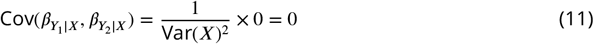

Since the covariance is zero, 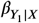 and 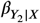 are uncorrelated. Given that *Y*_1_ and *Y*_2_ are independent, and 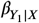 and 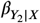 are derived from these independent variables, it follows that 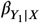 and 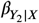 are independent.

## Appendix 2. Uniqueness of Value Ordering in Vectors from Cross Products

Given:

- A known vector *Y* of length *n*.
- Known vectors *X*_1_, *X*_2_, …, *X*_*m*_ of length *n*, but the order of the values in each *X*_*i*_ is unknown.
- Cross products *Y* ^′^*X*_1_, *Y* ^′^*X*_2_, …, *Y* ^′^*X*_*m*_.

We aim to show that for sufficiently large *m*, the ordering of the values in each vector *X*_1_, *X*_2_, …, *X*_*m*_ can be uniquely determined. Let *X*_*i*_ be the vector with unknown order, i.e., *X*_*i*_ = (*X*_*i*1_, *X*_*i*2_, …, *X*_*in*_). Let *σ*_*i*_ be a permutation of {1, 2, …, *n*} representing the unknown order of *X*_*i*_, so 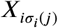 represents the *j*-th element of the permuted *X*_*i*_. The cross product 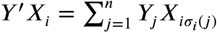 is given by different vectors *X*_1_, *X*_2_, …, *X*_*m*_, we have:

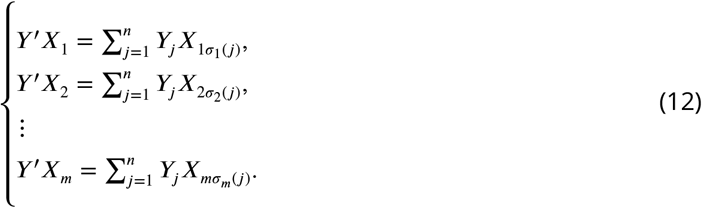

Let **Y** be the diagonal matrix with elements of *Y* on the diagonal, i.e., **Y** = diag(*Y*_1_, *Y*_2_, …, *Y*_*n*_). Let **X** be the *n* × *m* matrix with columns *X*_1_, *X*_2_, …, *X*_*m*_. The cross products can be represented as:

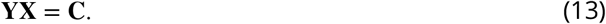

where **C** is an *n* × *m* matrix with each element *C*_*ij*_ = *Y*_*i*_*X*_*ij*_.

To show uniqueness, consider the matrix **A** formed by permuting the columns of **X** according to *σ*_*i*_:

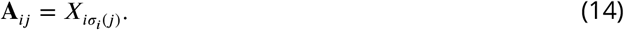

For sufficiently large *m*, the number of equations (cross products) *Y* ^′^*X*_*i*_ exceeds the number of unknowns (permutations *σ*_*i*_). Specifically, if *m > n*!, the system of equations becomes over-determined, meaning there are more constraints than possible permutations. Assume the vectors *X*_1_, *X*_2_, …, *X*_*m*_ are linearly independent. The matrix **X** has full rank, implying that the permutations *σ*_*i*_ can be uniquely determined by the cross products.

Since the system of equations *Y* ^′^*X*_*i*_ is over-determined and the vectors *X*_*i*_ are linearly independent, the permutations *σ*_*i*_ that reorder the elements of *X*_*i*_ must be unique. Any other permutation would result in a different set of cross-products, which contradicts the given *Y* ^′^*X*_*i*_. Therefore, for sufficiently large *m*, the cross products *Y* ^′^*X*_1_ to *Y* ^′^*X*_*m*_ uniquely determine the ordering of the values in each of the vectors *X*_1_, *X*_2_, …, *X*_*m*_. This completes the proof.

